# KNOWLEDGE AND AWARENESS OF OSTEOPOROSIS AMONG 144 YOUNG WOMEN AT CALMETTE HOSPITAL

**DOI:** 10.1101/2022.05.24.22275512

**Authors:** SUM Satha, UY Putthimundul

## Abstract

**Objectives:** To evaluate the level of knowledge and awareness about osteoporosis of young women at Calmette hospital.

**Methods:** We conveniently selected 144 women based on the selecting criteria. They were interviewed in Khmer language, face-to-face using structured-questionnaire which include demographic characteristic, lifestyle information, knowledge of osteoporosis and interest in an osteoporosis prevention.

**Results:** The average age of the participants was 28.43 (±4.42) years. Self-employed represented a large proportion. The study showed that only one fourth of participants went to a higher education. Most of the participants stated that they have known about osteoporosis while friends or family was the most common source of information. Only 16.67% have known osteoporotic patient. Most of them have a moderately bone-friendly lifestyle. Their calcium source depended largely on green vegetable. The average mean score of knowledge was 9.34 (±3.08) (range from 0 to 20). However, a half of them worried about suffering of osteoporosis in their later years of life. Around two thirds wanted to undergo an osteoporosis screening test. A majority was interested in osteoporosis prevention. Social media and community outreach were recommended as sensitizing measures. The osteoporosis knowledge was only correlated significantly (P-value: 0.007) with educational level.

**Conclusion:** Despite of low level of knowledge of osteoporosis, young Cambodian women was interested in osteoporosis screening and prevention. Educational intervention, such as public awareness programs regarding a bone-friendly lifestyle and osteoporosis screening, is crucial in sensitizing the general population about osteoporosis and its complication.

## Introduction

Osteoporosis might sound like an old term, but it is actually not, since it was invented and firstly described in early the 18^th^ century. It is a result of a combination of 2 Greek words: “ostéon” meaning bone and “poros” meaning passage or little hole. Based on these, osteoporosis means that bone with little holes [1]. Osteoporosis was linked to postmenopausal status until early 1940s and with calcium deficiency in the 1960s.

According to the American journal of Medicine, osteoporosis is a disease characterized by low bone mass and micro architectural deterioration of bone tissue, leading to enhanced bone fragility and a consequent increase in fracture risk [2]. It is a non-communicable disease and asymptomatic; a silent bone-fracturing disease; until its complications; osteoporotic fractures; occur [3]

The prevalence of osteoporosis is about 15% among the women aged 50 and over, and it increases to 80% for those over the age of 80 [4]. By 50 years of age, one in three women and one in five men will suffer a fracture in their remaining lifetime [5].

Osteoporosis has the physical and psychological impacts on the patient since 40% of survived fragility fracture patients become dependent and 60% require assistance a year later [6]. According to the International Osteoporosis Foundation, osteoporosis makes the patients to be hospitalized for more days in comparison with other diseases like breast cancer, myocardial infarction or diabetes [7]. Osteoporosis and fracture disrupt the global economic badly as they cost over 70 billion dollars in the first year following hip fractures [8].

As the prevalence of osteoporosis and its complications in South-East Asia countries, as in the global range, increase exponentially [9,10], we can assume that the number of osteoporosis in Cambodia increase as well even there are not any studies about Cambodia osteoporosis prevalence. It is a huge burden for the society.

The knowledge about osteoporosis can helps to prevent and to minimize the complications such as fragility fracture. Raising the knowledge of young women is then considered as a part of osteoporosis primary prevention [11,12].

Up until now there has been no data concerning osteoporosis knowledge in Cambodia. Therefore, this study is conducted in order to evaluate the knowledge and awareness about osteoporosis among young women in Cambodia that will be used to organize effective education programs.

## Methods

### Study setting and population

A prospective cross-sectional study was carried out in General Medicine “A” Department of Calmette hospital from 1^st^ July to 30^th^ September 2021. Out of 1314 attendants, after ruling out those whose consciousness is decreased, whose general appearance is unstable and who are diagnosed with or whose chief complaint is concerned with musculoskeletal disorder, there are 144 women aged from 20 to 35, and they were conveniently selected. Each participant provided written informed consent to this study before undertaking a face-to-face interview without tape recording. This study was permitted by University of Health Sciences and Calmette Hospital and also approved by the National Ethics Committee for Health Research of Cambodia.

### Study measure and data collection

A structured-questionnaire (Annexes 1) was used to collect the data. There are four main parts including: demographic characteristic, lifestyle information, knowledge of osteoporosis and interest in an osteoporosis prevention.

Demographic characteristics include age (years), residence (capital and province), occupation, educational level (illiteracy, primary school, high school, bachelor, master or PhD). For lifestyle information, we ask about daily exercise (no exercise or less than 90 minutes per week or more than or equal to 90 minutes per week), alcohol consumption (no consumption or less than 14 glasses per week or more than or equal to 14 glasses per week), smoking (yes/no) and calcium rich food consummation including milk (no consumption or less than 2 glasses per day or more than or equal to 2 glasses per day) and green vegetables (yes/no).

The knowledge about osteoporosis is evaluated by 20-item instrument addressing the generality of osteoporosis, risk factors, preventive methods and treatment. This instrument is derived from Osteoporosis Knowledge Assessment Tool (OKAT) to adapt to actual situation in Cambodia. The OKAT was translated from English to Khmer by a person proficient in both languages. This version was assessed by a senior Endocrinologist and a Professor of Public Health. It was piloted among 10 patient’s care givers at Calmette hospital to evaluate the suitability and coherence of content that leaded to few modifications of the questionnaire to suit the Cambodian context. Knowledge score are created by assigning a “1” to each correct response and a “0” to each incorrect or unknown. The items are summed for a possible range of 0 to 20, with higher scores indicating greater knowledge [13]. Along with OKAT questionnaire, the respondents were asked about which sex is at risk of osteoporosis and the consequence of monosodium glutamate consummation.

Regarding interest about osteoporosis, the participants were asked about their intention in osteoporosis screening test and whether they have searched about osteoporosis information and ever thought of risking of osteoporosis in the future.

### Statistical analysis

The data were entered to Excel 2016 and double-checked to avoid the errors. They were then exported to STATA version 14.2 (Stata Corp, College Station, Texas) to analyze statistically. Descriptive analyses were conducted for each variable to calculate frequency and proportion (%) for categorical variables and mean and standard deviation (SD) for continuous variables. Multiple linear regression was performed to assess the association between patient’s characteristics and knowledge score on osteoporosis. A p-value < 0.05 was considered statistically significant.

## Results

### Demographic characteristic of the participants

In total, 144 women at Calmette hospital were enrolled to the survey. The average age of the participants was 28.43 (±4.42) years (age range, 20 to 35 years). Self-employed represented about one fifth (21.53%) of the participant, followed by private office worker (18.06%), housewife (16.67%), factory worker (15.97%) and public officer (9.72%). Approximately 40% of the participants were from Phnom Penh, while just under two thirds were originated from provinces, particularly Kandal and Kompong Cham. Two thirds of the participants were married, while divorce only represented just under 5%. The study showed that approximately 50% finished high school. Only 5 participants completed post bachelor degree, while another 5 are illiterate and all of them came from province.

**Table 1.** Demographic characteristics

| Variables | Frequency | Percentage |
| --- | --- | --- |
| **Age in years, mean (±SD)** | 28.43 ($\pm 4.42$) | |
| < 25 | 29 | 20.14 |
| 25 – 29 | 56 | 38.89 |
| $\geq 30$ | 59 | 30.97 |
| **Professions** |  |  |
| Factory workers | 23 | 15.97 |
| Farmers | 15 | 10.42 |
| Housewife | 24 | 16.67 |
| Private office worker | 26 | 18.06 |
| Public officer | 14 | 9.72 |
| Self-employed | 31 | 21.53 |
| Student | 9 | 6.25 |
| Other | 2 | 1.39 |
| **Current address** |  |  |
| Phnom Penh | 58 | 40.28 |
| Province | 86 | 59.72 |
| **Marital status** |  |  |
| Single | 41 | 28.47 |
| Married | 96 | 66.67 |
| Divorce | 7 | 4.86 |
| **Level of education** |  |  |
| Illiteracy | 5 | 3.47 |
| Primary school | 34 | 23.61 |
| High school | 65 | 45.14 |
| Bachelor degree | 35 | 24.31 |
| Post bachelor degree | 5 | 3.47 |

### Source of osteoporosis information

Most of the participants (98.1%) stated that they have known about osteoporosis. Most of them (58.33%) have heard of osteoporosis from their friends or family. Social media and physician were second source of information, each represents 15.28%. Only 16.67% have known osteoporotic patient.

**Table 2.**
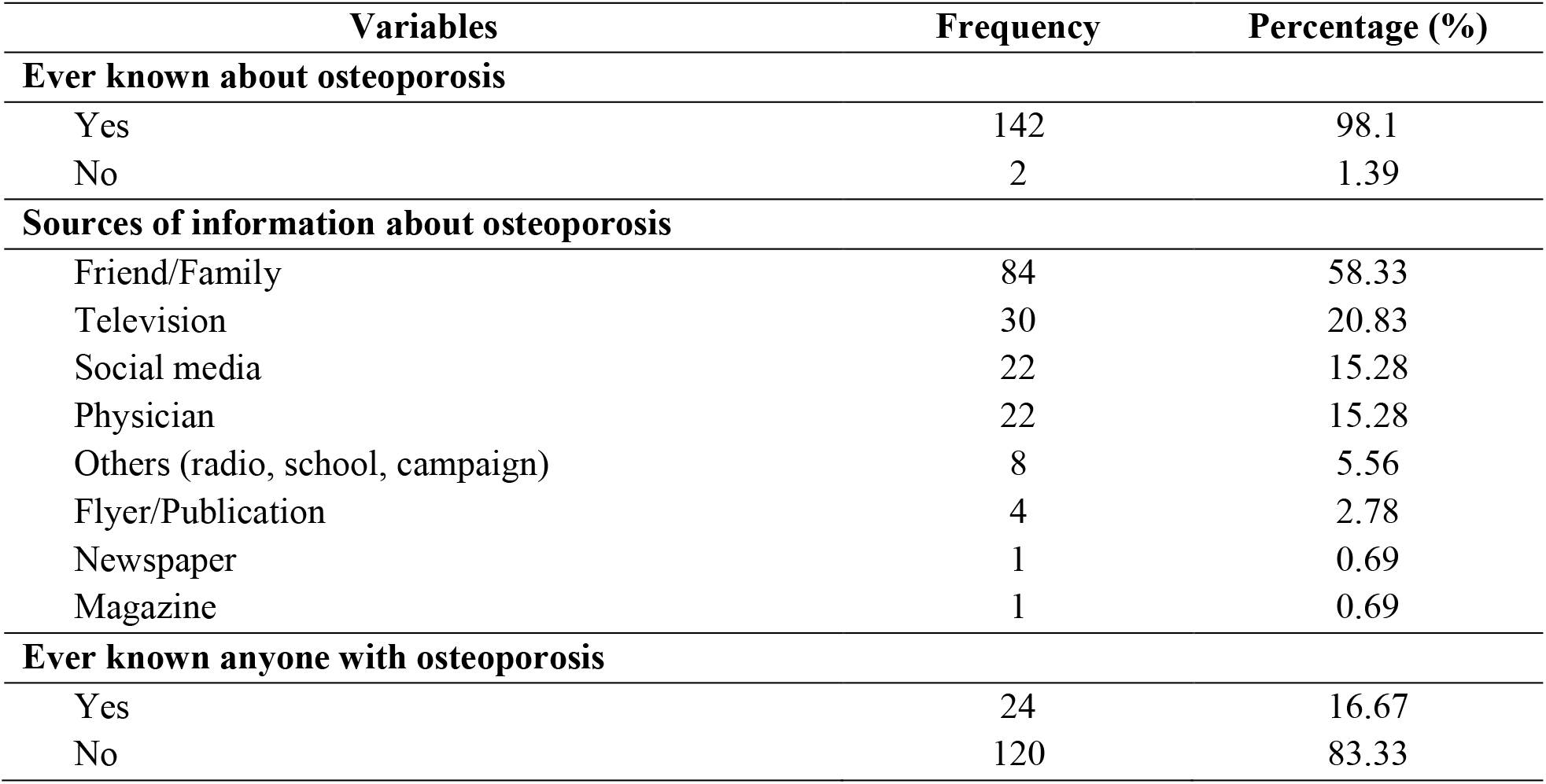
Source of information about osteoporosis

| **Variables** | **Frequency** | **Percentage (%)** |
| --- | --- | --- |
| **Ever known about osteoporosis** |  |  |
| Yes | 142 | 98.1 |
| No | 2 | 1.39 |
| **Sources of information about osteoporosis** |  |  |
| Friend/Family | 84 | 58.33 |
| Television | 30 | 20.83 |
| Social media | 22 | 15.28 |
| Physician | 22 | 15.28 |
| Others (radio, school, campaign) | 8 | 5.56 |
| Flyer/Publication | 4 | 2.78 |
| Newspaper | 1 | 0.69 |
| Magazine | 1 | 0.69 |
| **Ever known anyone with osteoporosis** |  |  |
| Yes | 24 | 16.67 |
| No | 120 | 83.33 |

### Lifestyle of the participants

About 60% of the participants did not exercise (63.89%) regularly nor drink milk daily (57.64%). Four quarters consumed green vegetable daily. None of them smoked. Only 10% of them drank alcohol occasionally.

**Table 3.** Lifestyle of respondents

| Variables | Frequency | Percentage (%) |
| --- | --- | --- |
| **Exercise** |  |  |
| No | 92 | 63.89 |
| < 90 minute per week | 25 | 17.36 |
| ≥ 90 minutes per week | 27 | 18.75 |
| **Milk consumption per day** |  |  |
| No | 83 | 57.64 |
| < 2 glasses per day | 49 | 34.03 |
| ≥ 2 glasses per day | 12 | 8.33 |
| **Daily green vegetable consumption** |  |  |
| No | 33 | 22.92 |
| Yes | 111 | 77.08 |
| **Cigarette smoking** |  |  |
| No | 144 | 100.00 |
| Yes | 0 | 0.00 |
| **Alcohol drinking** |  |  |
| No | 131 | 90.97 |
| < 14 glasses per week | 12 | 8.33 |
| ≥ 14 glasses per week | 1 | 0.69 |

### Knowledge and awareness level

The average mean score of knowledge was 9.34 (±3.08) (range from 0 to 20). Among 20 statements of OKAT-derived questionnaire, the 13^th^ phrase: “An adequate calcium intake can be achieved from two glasses of milk a day.” was the question that was answered correctly the most, when the 2^nd^ phrase: “2. Osteoporosis usually causes symptoms (e.g. pain) before fractures occur.” the least.

A large majority of the participants (84.72%) misbelieved that monosodium glutamate consummation leads to osteoporosis. Approximate one third (36.11%) of the respondents correctly knew that women were at risk of osteoporosis. Just under 38.19% of the participants thought that both male and female were equally at risk. Moreover, around 10% believed that men were at more risk than women.

**Table 4.** Knowledge on osteoporosis

| OKAT questionnaire | Correct response |  |
| --- | --- | --- |
|  | Frequency | Percentage (%) |
| 1. Osteoporosis leads to an increased risk of bone fractures. | 110 | 76.39 |
| 2. Osteoporosis usually causes symptoms (e.g. pain) before fractures occur. | 10 | 6.94 |
| 3. Having a higher peak bone mass at the end of childhood gives no protection against the development of osteoporosis in later life. | 60 | 41.67 |
| 4. Osteoporosis is more common in men. | 106 | 73.61 |
| 5. Cigarette smoking can contribute to osteoporosis. | 68 | 47.22 |
| 6. White women are at highest risk of fracture as compared to other races. | 26 | 18.06 |
| 7. A fall is just as important as low bone strength in causing fractures. | 112 | 77.78 |
| 8. By age 80, the majority of women have osteoporosis. | 106 | 73.61 |
| 9. From age 50, most women can expect at least one fracture before they die. | 70 | 48.61 |
| 10. Any type of physical activity is beneficial for osteoporosis. | 22 | 15.28 |
| 11. It is easy to tell whether I am at risk of osteoporosis by my clinical risk factors. | 56 | 38.89 |
| 12. Family history of osteoporosis strongly predisposes a person to osteoporosis. | 37 | 25.69 |
| 13. An adequate calcium intake can be achieved from two glasses of milk a day. | 116 | 80.56 |
| 14. Sardines and broccoli are good sources of calcium for people who cannot take dairy products. | 38 | 26.39 |
| 15. Calcium supplements alone can prevent bone loss. | 97 | 67.36 |
| 16. Alcohol in moderation has little effect on osteoporosis. | 60 | 41.67 |
| 17. A high salt intake is a risk factor for osteoporosis. | 68 | 47.22 |
| 18. There is a small amount of bone loss in the 10 years following the onset of menopause. | 63 | 43.75 |
| 19. Hormone therapy prevents further bone loss at any age after menopause. | 42 | 29.17 |
| 20. There are no effective treatments for osteoporosis available in “Cambodia”. | 78 | 54.17 |

### Interest of the participant in osteoporosis

Most of the participants (92.36%) claimed that they never searched about osteoporosis before, but a half of them (52.18%) of them were afraid of suffering of osteoporosis in their elder hood. Around two thirds (69.45%) wanted to undergoing an osteoporosis screening test. Furthermore, a majority (95.13%) were interested in osteoporosis prevention. The study also found that social media (64.58%), Facebook particularly mentioned, and community outreach (34.02%) were recommended for sensitizing the general population about osteoporosis and preventive measures.

**Table 5.**
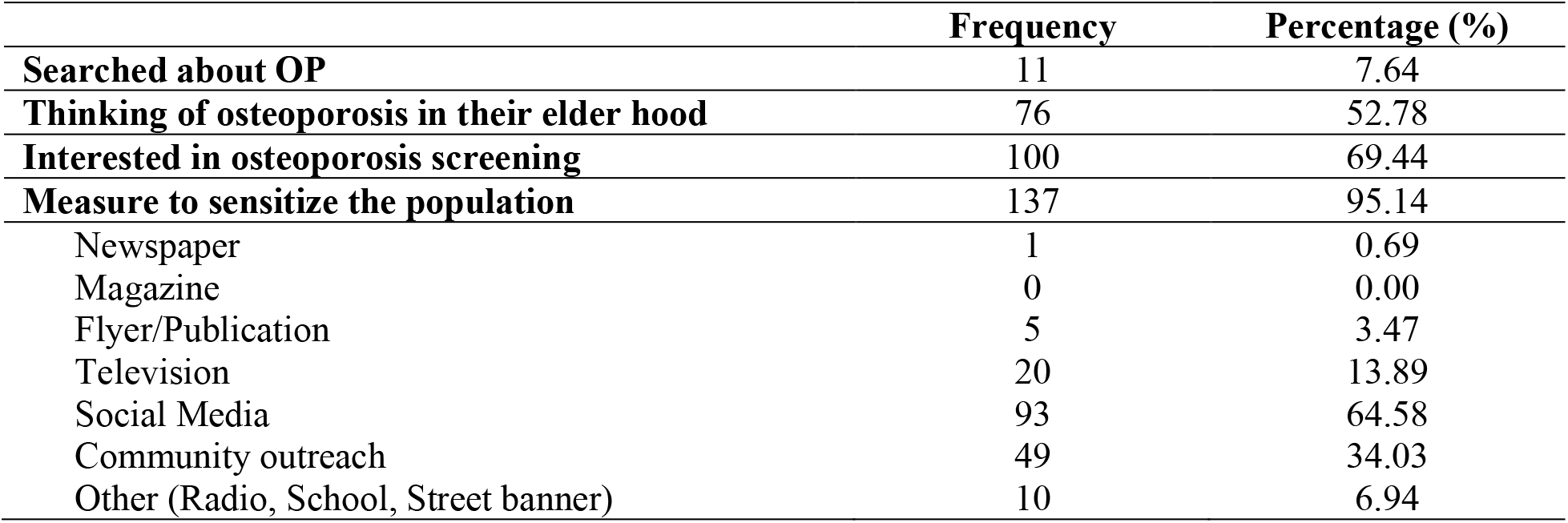
Interest in osteoporosis

### Association between osteoporosis knowledge level and demographic characteristic

All factors considered, the osteoporosis knowledge was only correlated significantly (P-value: 0.007) with educational level. Specifically, the participants who completed post bachelor degree have statistically greater knowledge score, in comparison with who never go to school.

**Table 6.** The associations between risk factors and overall awareness score in the multivariable linear regression models

| Variables | $\beta$ | SE | P-value |
| --- | --- | --- | --- |
| **Age in years** |  |  |  |
| 25-29 vs. < 25 | - 0.936 | 0.855 | 0.276 |
| $\geq 30$ vs. < 25 | - 0.937 | 0.852 | 0.274 |
| **Province vs. Phnom Penh** | - 0.277 | 0.636 | 0.664 |
| **Occupation** |  |  |  |
| Farmers vs. factory workers | 1.072 | 1.098 | 0.331 |
| Housewives vs. factory workers | - 0.807 | 0.935 | 0.390 |
| Others vs. factory workers | - 0.326 | 2.567 | 0.899 |
| Private office workers vs. factory workers | - 0.546 | 1.085 | 0.615 |
| Public officers vs. factory workers | 0.802 | 1.232 | 0.516 |
| Self-employed vs. factory workers | 0.311 | 0.928 | 0.738 |
| Students vs. factory workers | 0.893 | 1.556 | 0.567 |
| **Marital status** |  |  |  |
| Married vs. single | 0.152 | 0.769 | 0.844 |
| Divorce vs. single | 0.458 | 1.397 | 0.741 |
| **Level of education** |  |  |  |
| Primary school vs. no school | 1.022 | 1.603 | 0.525 |
| High school vs. no school | 1.017 | 1.603 | 0.527 |
| Bachelor degree vs. no school | 1.973 | 1.743 | 0.260 |
| Post bachelor degree vs. no school | 6.225 | 2.259 | **0.007** |
| **Exercise** |  |  |  |
| < 90 minute per week vs. none | 0.854 | 0.756 | 0.261 |
| $\geq 90$ minutes per week vs. none | 0.333 | 0.780 | 0.670 |
| **Milk consumption per day** |  |  |  |
| < 2 glasses per day vs. none | 0.691 | 0.626 | 0.272 |
| $\geq 2$ glasses per day vs. none | - 0.141 | 1.059 | 0.895 |
| **Daily green vegetable consumption vs. no** | - 0.281 | 0.646 | 0.665 |
| **Cigarette smoking vs. no** | N/A |  |  |
| **Alcohol drinking** |  |  |  |
| < 14 glasses per week vs. no | 0.259 | 1.042 | 0.804 |
| $\geq 14$ glasses per week vs. no | 4.427 | 3.272 | 0.179 |
| **Sources of information about osteoporosis** |  |  |  |
| Newspaper | 0.677 | 3.385 | 0.984 |
| Magazine | 2.422 | 3.710 | 0.515 |
| Flyer/Publication | 0.704 | 1.771 | 0.692 |
| Television | - 0.077 | 0.808 | 0.923 |
| Social media | 1.379 | 0.931 | 0.142 |
| Friend/Family | 0.091 | 0.863 | 0.916 |
| Physician | 0.029 | 0.944 | 0.976 |
| Others (radio, school, campaign) | - 0.772 | 1.318 | 0.559 |
| **Ever known anyone with osteoporosis vs. no** | 0.640 | 0.811 | 0.432 |

Abbreviations: $\beta$ = regression coefficient; SE = standard error

## Discussion

This study evaluated the knowledge and awareness of osteoporosis of young Cambodian women who attended General Medicine A ward of Calmette Hospital. It restricted participants aged between 20 and 35 years in order to understand the basic information and the current level of knowledge and awareness of osteoporosis among young women that could be used for raising awareness and taking public health measures in the prevention and prediction of osteoporosis and its complications within 10 to 25 years later.

One hundred and forty-two women (98.1%) in our survey had heard about osteoporosis, mostly through their friends or family (58.33%). While in the study conducted in Thailand and China, more than half of participants reported television as their main source of osteoporosis knowledge [14,15].

It seems that media in Cambodia, including newspapers, magazine, television and social media, does not provide enough health news to the general population since it represented less than 50% of the source of information. Either Cambodian population do not have enough access to media.

Only around fifteen percent of them obtained the information from a health care provider. It is comparable to a study conducted in Thailand [14]. It might signify that health care providers did not give osteoporosis information to their patients or the information was not correctly received. It looked like the crucial role of practitioners to cure their patients but to not give a preventive care.

Cambodian national education program did not seem to give attention to raising awareness about osteoporosis since only 3 women (2.08%) have marked school as their source of information. Neither primary school nor high school was significantly correlated to level of knowledge. Additionally, even the women who are obtaining bachelor degree did not have sufficient knowledge about osteoporosis. Only the women that completed post bachelor degree had a great osteoporosis knowledge (P-value = 0.007). In Vietnam population, women who finished high school had a great knowledge about OP [16]. This result was no different from that in a Chinese population, where the study included both male and female of any age [15].

Our study found that young Cambodian women were interested in getting information from social networking, particularly Facebook. In addition, direct community outreach shared an essential role in the osteoporosis raising awareness campaign. It is different from the other studies where television was considered as an important and effective measure to disseminate the osteoporosis knowledge to the population [14-16].

120 out of 144 attendants (83.33%) have never known anyone with osteoporosis while the its prevalence was high among South-East Asia countries, as in the global range [9,10]. It signified that the osteoporosis was overlooked in Cambodia. In actuality, there are only two DXA-scan machines available among around 10 tertiary national hospitals but the availability is higher at private sector.

Even though over 90% of attendants claimed that they have known about osteoporosis, most of them had less than adequate knowledge about osteoporosis as the mean score was 9.34 (±3.08). This was probably due to the low quality of information about osteoporosis available. For instance, the statement “Monosodium glutamate leads to osteoporosis” was believed as a fact by a majority (84.72%) despite having no supporting evidence whatsoever. It is confirmed that this misbelief was disseminating in Cambodia. Nevertheless, it does not mean that monosodium glutamate consumption is encouraged.

The overall average mean score (46.7%) in this study is comparable with a study in Vietnam (49%) while lower than the other studies (80.43% in Thailand and 67.8% in China) [14-16].

Osteoporosis risk factors were not well known among the attendants. Osteoporosis was not known as a stereotypical repercussion of cigarette smoking, alcohol consumption and high salt intake as less than average of the attendants answered correctly to the statement 5, 16 and 17 (47.22%, 41.67% and 47.22% respectively).

In case of statement 4: “Osteoporosis is more common in men”, 106 of 144 attendants (73.61%) correctly recognized it as false. However, only one third (36.11%) of the respondents correctly claim that women were at risk of osteoporosis. Therefore, OKAT questionnaire should be rectified because though the respondents select the correct answer (False), it did not signify they had a true concept. Additionally, most of them were not aware of their possible risk of osteoporosis in next 20 years.

Most of the respondents (77.08%) consumed green vegetable daily, while more than a half of them (57.64%) did not drink milk. These might ensue due to the availability and the price of green vegetable which were cheaper and more convenient than milk. Most of Cambodian women cannot drink milk due to different reasons. Additionally, most of the milk at Cambodia market were imported from abroad and the population preferred the fresh and free of preservative ingredient. The total of participants (100%) did not smoke and only less than 10% drank alcohol. These may be explained by the cliché that smoking among women is a sign of bad image. It is also a result of forceful and effective Cambodian National Campaign of Ministry of Health against smoking in public and cigarette advertising. However, we observed that more and more women drank alcohol socially.

Since our study is the first study about osteoporosis conducted in Cambodia, there are two limitations. Firstly, it is carried out in a single center and is not a population-based. Secondly, this study included only 144 women; a small sample size has limitation in representing the general Cambodian population. Last but not least, our study design is different from the other studies because of either the questionnaire, or sample population. Therefore, the comparison is restricted to the overall view.

## Conclusion

In short, our survey based on cross-sectional study revealed a low knowledge and awareness level about osteoporosis among young women in this study, except for those who finished post bachelor degree. Most of them had a moderately bone-friendly lifestyle as they did not smoke, nor drink. Their calcium source depended largely on green vegetable. The rate of women who drink milk regularly is low. However, they were interested in osteoporosis screening test and prevention after having had been informed of their complications.

## Data Availability

All data produced in the present work are contained in the manuscript.

## Acknowledgement

We would like to specially thank to Mr. AN YOM for advices in statistical analyzing.

